# Efficacy and Safety of Nasal High-Frequency Oscillation in Preventing Intubation in Very-Low-Birth-Weight Infants with Respiratory Distress Syndrome

**DOI:** 10.1101/2024.05.10.24307155

**Authors:** Buu Quoc Dang, Tam Thi Thanh Pham, Duc Ninh Nguyen, Nguyen Phuoc Long, Tinh Thu Nguyen

## Abstract

**Background:** Invasive mechanical ventilation in very-low-birth-weight infants (VLBWI) was associated with immediate and long-term complications. Nasal high-frequency oscillation (nHFO) has recently become a new non-invasive ventilation (NIV) mode for treating respiratory failure in VLBWI. This study aimed to investigate the safety and efficacy of nHFO as an alternative respiratory support to prevent intubation in VLBWI.

**Methods:** A retrospective analysis was conducted using the clinical data of 42 VLBWIs with respiratory distress syndrome (RDS) who were treated in our department from August 2018 to August 2020 and met the selection criteria.

**Results:** nHFO was used as a rescue strategy in 32 infants and a prophylactic strategy in 10 infants. It was observed that out of 42 cases, 30 cases (71.4%) were able to avoid intubation within 72 hours, while 23 cases (54.8%) were successfully switched to another NIV mode from nHFO. There was a significant decrease in pCO2 and an increase in pH one hour after using nHFO in the success group. Two cases (4.8%) of feeding intolerance associated with nHFO were noted.

**Conclusion:** This study showed that nHFO as alternative respiratory support for preterm infants with RDS might be safe and effective in reducing the need for intubation.

**Highlights:**

i. Invasive mechanical ventilation in very low-birth-weight infants (VLBWI) was associated with immediate and long-term complications. As a result, there is an increasing focus on researching and applying non-invasive ventilation (NIV).
ii. In this study, Nasal high-frequency oscillation (nHFO) in VLBWI with respiratory distress syndrome showed promising efficacy and safety, with 71.4% of cases avoiding intubation within 72 hours. Further, 54.8% of cases were successfully transitioned to another NIV mode from nHFO.
iii. Further randomized controlled trials with adequately powered sample sizes are warranted to assess the overall safety and efficacy of nHFO.

## Introduction

Every year, approximately 13.4 million infants are born preterm (born less than 37 weeks of gestational age), representing a prevalence rate of 4-16% of total live births globally ^1^. The implications of preterm birth are profound, as complications arising from it account for 35% of all neonatal mortalities and 17.7% of all deaths in children under the age of five years ^2^. A significant morbidity observed in infants born with very low birth weight (VLBW) is respiratory distress syndrome (RDS), primarily attributable to the inadequate maturation of the pulmonary system at the time of birth. The prevalence and severity of RDS in this population highlight the critical need for advancements in neonatal care and interventions aimed at addressing the challenges associated with preterm births.

At our neonatal intensive care unit (NICU), Children’s Hospital 1, Ho Chi Minh City, Vietnam), we annually manage around 700 VLBW preterm infants requiring intensive care for prematurity-related complications. All admitted infants experience varying degrees of respiratory failure, with 24.4% necessitating endotracheal intubation on admission ^3^.

Invasive mechanical ventilation in neonates is known as a risk factor for later development of other morbidities, including bronchopulmonary dysplasia and intraventricular hemorrhage ^4-6^. Non-invasive ventilation (NIV) use has been suggested to mitigate those complications. Nasal high-frequency oscillation (nHFO) has emerged as a promising NIV strategy for neonates. Recent studies suggested its advantages in preventing the need for invasive mechanical ventilation ^7,8^. Since 2018, our local unit has been using nHFO for preterm infants who have failed other NIV modes, such as nasal continuous positive airway pressure - nCPAP, biphasic positive airway pressure - BIPAP, and nasal intermittent positive pressure ventilation - NIPPV. Several lines of evidence suggest that nHFO may be more effective than nCPAP and other NIV modalities in preventing extubation failure in preterm neonates ^9,10^. Other studies have focused on the effectiveness of nHFO after the failure of other NIV modes and have shown promising results ^11^.

This study aimed to investigate the safety and efficacy of nHFO as an alternative respiratory support to prevent intubation in VLBWI.

## Methods

### Ethical approval and study design

We conducted a retrospective study by analyzing the clinical data of all premature infants with RDS who were treated with nHFO in the NICU at Children’s Hospital 1, Ho Chi Minh City, Vietnam, from August 2018 to August 2020. The study was approved by the Institutional Review Board (IRB) of Children’s Hospital 1 (IORG0007285, FWA00009748) on April 7, 2022 (Project No: CS/N1/22/10). A waiver of informed consent was granted by the IRB due to the retrospective nature of the study. Patient data was anonymized and de-identified throughout the analysis. Despite a shift towards an approach of pre-emptive treatment with surfactant based on clinical assessment of breathing patterns and inspired oxygen requirements to avoid worsening RDS, our study employed established clinical criteria for RDS diagnosis in all participants. These criteria included the combination of non-specific respiratory symptoms (tachypnea, nasal flaring, grunting, retractions, and cyanosis) and chest radiography findings (homogenous lung disease with diffuse atelectasis, ground-glass reticulo-granular appearance with air bronchograms, and low lung volumes) ^12^. In our unit, all cases on NIV modes were treated with surfactant replacement therapy using the less-invasive surfactant administration (LISA) method. Infants who were intubated received surfactant via an endotracheal tube, and efforts were made to extubate them to NIV modes as early as possible. The ventilators used in our unit do not support synchronization with the patient’s breathing during non-invasive ventilation (NIV).

### Setting and Participants

The retrospective analysis was conducted at the NICU of Children’s Hospital 1, Ho Chi Minh City, Vietnam. The inclusive criteria were as follows: (i) premature infants with gestational age < 32 weeks and birth weight < 1500 grams; (ii) diagnosed with RDS; and (iii) unresponsive to other NIV modes and indication of invasive mechanical ventilation or weaned from invasive mechanical ventilation with a high risk of reintubation. The exclusion criteria encompassed: (i) pulmonary hemorrhage; (ii) complications involving severe congenital anomaly, including congenital heart disease, congenital diaphragmatic hernia, respiratory tract malformation, and severe digestive tract malformation; (iii) preoperative status; (iv) incomplete data.

### Interventions

There were two indications of nHFO use: rescue, when the infant transitioned from another NIV mode, or prophylactic, when the infant was weaned from invasive mechanical ventilation. The efficacy of nHFO at two specific time points was evaluated: (i) within 72 hours after initiating nHFO, infants who avoided intubation were categorized as the “72-hour success group”; the remainder was termed “72-hour failure group”; (ii) at the completion of nHFO, infants who successfully transitioned to another NIV mode were categorized as the “final success group”, whereas the rest were designated as the “final failure group”.The indications for initiating intubated ventilation in our unit^13,14^ included (i) deterioration of respiratory function following NIV (based on one of the following clinical signs: (1) increased work of breathing such as tachypnea, use of accessory muscles, nasal flaring, grunting, or retractions, (2) hemodynamic instability such as changes in blood pressure and heart rate, or (3) decreased consciousness or lethargy), (ii) severe apnea characterized by more than three episodes per hour and a heart rate below 100 beats per minute or requiring bag-and-mask ventilation, (iii) pulmonary hemorrhage, (iv) severe respiratory acidosis, defined as a PCO2 level exceeding 65 mmHg and a pH lower than 7.2, or (v) severe hypoxemia, where the FiO2 exceeds 0.5, accompanied by either a PaO2 level below 45 mmHg or a SpO2 level below 90%.

### Instruments

nHFO was provided by using either a Babylog 8000 plus ventilator (Dräger, Lübeck, Germany) or Sensor Medics 3100A ventilator (CareFusion, California, USA) via a RAM cannula (Neotech, California, USA). An orogastric tube was kept open to decompress the stomach and to facilitate feeding.

Initial settings for MAP and frequency on nHFO were 12 to 18 cmH2O and 8 to 11 Hz, respectively. Amplitude ranged from 35 to 45 cmH2O on the Sensor Medics 3100A and started with 100% on the Babylog 8000 plus. Adjustments were guided by blood gas analysis and peripheral oxygen saturation (SpO2) measurements. FiO2 was adjusted in increments of 0.05, MAP by 1 cmH2O, and frequency by 1 Hz each time.

### Outcomes

The primary outcomes of the study included the success rate of avoiding intubation, including (i) within 72 hours after starting nHFO, patients who required intubation were classified as the “72h failure group” (the rate of this group is called the “failure rate 72h”), while those who were still on nHFO or transitioned to another non-invasive ventilation method were classified as the “72h success group” (the rate of this group is called the “success rate 72h”); (ii) After 72 hours, patients who required intubation were classified as the “final failure group” (the rate of this group is called the “failure rate final”), and those who transitioned to another non-invasive ventilation method were classified as the “final success group” (including those who transitioned to another non-invasive ventilation method within the first 72 hours) (the rate of this group is called the “success rate final”).

The secondary outcomes of the study were blood gas parameters before and after the application of nHFO (pH, pCO_2_, pO_2_, and BE) and complications associated with nHFO use, including necrotizing enterocolitis (NEC), intestinal perforation, intra-ventricular hemorrhage (IVH), nasal columella injury, and feeding intolerance.

Diagnosis of NEC was based on Bell’s criteria ^15^, intestinal perforation determined by the sign of free abdominal air on an x-ray or by exploration of a perforation in the gastrointestinal tract during surgery. IVH was determined according to Papile’s grade ≥ II ^16^ on ultrasound; nasal columella injury was determined based on the classification of Buettiker et al. ^17^. Feeding intolerance was defined as difficulty to digest enteral feedings and is accompanied by an increase in gastric residuals, abdominal distension, and/or reflux ^18^.

### Statistical Analysis

The analysis of the data was conducted with IBM SPSS Statistics (Version 22.0, IBM Corp.). The Shapiro-Wilks test was utilized to assess the distribution of continuous variables. Normally distributed values were analyzed using the Student’s t-test, while the Mann-Whitney U-test was applied to independent samples with non-normally distributed values. For a single sample with non-normally distributed values, the Wilcoxon matched-pairs test was utilized. Categorical variables were compared by using the Chi-square test. The level of statistical significance was set at p=0.05 for all tests.

## Results

Forty-three neonates were enrolled in the study. nHFO was employed as a rescue strategy in 33 of the 43 cases, while the remaining 10 received it as a prophylactic strategy. One infant in the rescue group was excluded from the analysis due to intubation within 72 hours for rescue surfactant replacement therapy. The baseline characteristics of all participants are presented in **Table 1**. A statistically significant difference was observed only in the rate of surfactant therapy (p < 0.05). All other characteristics were comparable between the two groups.

**Table 1.** Baseline characteristics.

| Characteristics | Total<br>(N = 42) | 72-hour success<sup>*</sup><br>(n = 30) | 72-hour failure<sup>†</sup><br>(n = 12) | p-value |
| --- | --- | --- | --- | --- |
| Male sex - n (%) | 17 (40.5) | 13 (43.3) | 4 (33.3) | 0.562 |
| Birth weight<sup>‡</sup> (gram) |  |  |  |  |
| Median (IQR) | 800 (650-900) | 800 (692-925) | 700 (600-830) | 0.068 |
| Gestational age<sup>‡</sup> (week) |  |  |  |  |
| Median (IQR) | 25 (25-27) | 25 (25-27.5) | 25 (24.3-25.8) | 0.240 |
| Weight at study entry<sup>‡</sup> (gram) |  |  |  |  |
| Median (IQR) | 888 (805-1130) | 995 (805-1227) | 860 (615-894) | 0.056 |
| PMA at study entry<sup>‡</sup> (week) |  |  |  |  |
| Median (IQR) | 29.1 (27.9-31.1) | 29.3 (28.3-32) | 28.5 (26.8-30.2) | 0.123 |
| Age at study entry<sup>‡</sup> (day) |  |  |  |  |
| Median (IQR) | 27 (14-34) | 28 (14-38) | 22 (13-30) | 0.189 |
| Surfactant therapy - n (%) | 34 (81) | 22 (73.3) | 12 (100) | < 0.001 |
| Antenatal steroids - n (%) |  |  |  |  |
| Complete | 14 (33.3) | 10 (33.3) | 4 (33.3) | 0.983 |
| Incomplete | 25 (59.6) | 18 (60) | 7 (58.3) |  |
| Unknown | 3 (7.1) | 2 (6.7) | 1 (8.3) |  |
|  |  | Final success<sup>§</sup><br>n = 23 | Final failure<sup>#</sup><br>n = 19 | p-value |
| Male - n (%) |  | 11 (47.8) | 13 (68.4) | 0.297 |
| Birth weight<sup>‡</sup> (gram) |  |  |  |  |
| Median (IQR) |  | 800 (700-900) | 700 (600-900) | 0.380 |
| Gestational age<sup>‡</sup> (week) |  |  |  |  |
| Median (IQR) |  | 25 (25-27.5) | 25 (25-26) | 0.676 |
| Weight at study entry<sup>‡</sup> (gram) |  |  |  |  |
| Median (IQR) |  | 1010 (810-1220) | 860 (790-980) | 0.160 |
| PMA at study entry<sup>‡</sup> (week) |  |  |  |  |
| Median (IQR) |  | 29.4 (27.8-32) | 29.1 (27.7-30.6) | 0.503 |
| Age at study entry<sup>‡</sup> (day) |  |  |  |  |
| Median (IQR) |  | 27 (14-35) | 27 (19-33) | 0.667 |
| Surfactant therapy - n (%) |  | 16 (69.6) | 18 (94.7) | 0.018 |
| Antenatal steroids - n (%) |  |  |  |  |
| Complete |  | 7 (30.4) | 7 (36.8) | 0.863 |
| Incomplete |  | 14 (60.9) | 11 (57.9) |  |
| Unknown |  | 2 (8.7) | 1 (5.3) |  |

Abbreviations: nHFO, Nasal High-Frequency Oscillation; PMA, post-menstrual age.
<sup>\*</sup>Infants who were precluded intubation within 72 hours after the initiation of nHFO; <sup>†</sup>infants who were intubated within 72 hours after the initiation of nHFO; <sup>‡</sup>results presented as median (IQR) and p values calculated using Mann Whitney U test; <sup>§</sup>infants who transitioned to another non-invasive mode at the completion of nHFO; <sup>#</sup>infants who were intubated at the completion of nHFO.

Overall, nHFO successfully averted 30 (71.4%) cases from intubation within 72 hours. At the completion of nHFO, 23 (54.8%) cases were successfully switched to another NIV mode (**Fig. 1**). The most frequent primary indication for nHFO initiation was spells (33%), followed by increasing work of breathing (24%), and hypercapnia (22%). Other less common indications (21%) included high FiO2 and difficult weaning from IMV/HFOV. The median duration of nHFO use was 72 hours (IQR: 41-124), with the most prolonged duration being 534 hours. There were no statistically significant differences in blood gas parameters before the use of nHFO between the 72-hour success group and 72-hour failure group (**Table 2**). Among neonates who successfully prevented intubation within 72 hours, there was a significant decrease in pCO2 and an increase in pH and SpO2 (all p values < 0.05) before and one hour after the use of nHFO (**Table 3**). On the other hand, the 72-hour failure group did not exhibit statistically significant changes in blood gas parameters one hour after nHFO use. Regarding complications associated with nHFO, two cases (4.8%) of feeding intolerance were documented, and no other adverse events were observed during nHFO.

**Table 2.** Changes in blood gas parameters before and one hour after the initiation of nHFO.

| Parameters | 72-hour success*<br>(n= 30) |  |  | 72-hour failure†<br>(n= 12) |  |  |
| --- | --- | --- | --- | --- | --- | --- |
|  | Pre | Post | P value | Pre | Post | P value |
| pH | 7.26 | 7.28 | 0.033 | 7.28 | 7.25 | 0.499 |
| Median (IQR) | (7.18-7.34) | (7.25-7.35) |  | (7.21-7.34) | (7.22-7.37) |  |
| pCO<sub>2</sub> (mmHg) |  |  | 0.028 |  |  | 0.128 |
| Median (IQR) | 46 (37-55) | 39 (34-47) |  | 42 (33-44) | 45 (37-56) |  |
| pO<sub>2</sub> (mmHg) |  |  | 0.758 |  |  | 0.866 |
| Median (IQR) | 56 (44-86) | 62 (47-78) |  | 54 (49-81) | 51 (45-69) |  |
| FiO<sub>2</sub> (%) |  |  | 0.179 |  |  | 0.858 |
| Median (IQR) | 40 (40-60) | 40 (38-50) |  | 50 (30-60) | 45 (33-70) |  |
| SpO<sub>2</sub> (%) |  |  | 0.016 |  |  | 0.136 |
| Median (IQR) | 93 (91-95) | 95 (93-96) |  | 92 (90-94) | 93 (92-95) |  |
| BE (mmol/L) |  |  | 0.970 |  |  | 0.062 |
| Median (IQR) | -7 (-9; -4) | -7 (-11; -4) |  | -8 (-10; -3) | -6 (-10; 0) |  |

Abbreviations: nHFO, Nasal High-Frequency Oscillation
\* Infants who were precluded intubation within 72 hours after nHFO use; † infants who were intubated within 72 hours after nHFO use; p-values calculated using Wilcoxon signed-rank test.

**Table 3.** Blood gas parameters before nHFO between 72-hour success and failure groups.

| Parameters | 72-hour success<sup>*</sup><br>(n= 30) | 72-hour failure<sup>†</sup><br>(n= 12) | P value |
| --- | --- | --- | --- |
| pH | 7.26 | 7.28 | 0.636 |
| Median (IQR) | (7.18-7.34) | (7.22-7.34) |  |
| pCO<sub>2</sub> (mmHg) | 46.1 | 41.7 | 0.328 |
| Median (IQR) | (36.8-54.8) | (34-44.1) |  |
| pO<sub>2</sub> (mmHg) | 55.5 | 54 | 0.901 |
| Median (IQR) | (44-79) | (51.5-79.5) |  |
| FiO<sub>2</sub> (%) | 40 | 50 | 0.914 |
| Median (IQR) | (40-60) | (35-60) |  |
| SpO<sub>2</sub> (%) | 93 | 92 | 0.179 |
| Median (IQR) | (91-95) | (90.5-93) |  |
| BE (mmol/L) | -7 | -8 | 0.784 |
| Median (IQR) | (-9; -4) | (-9.5; -4) |  |

Abbreviations: nHFO, Nasal High-Frequency Oscillation.
<sup>\*</sup> Infants who were precluded intubation within 72 hours after nHFO use; <sup>†</sup> infants who were intubated within 72 hours after nHFO use; p-values calculated using Mann-Whitney U test.

**Fig. 1.**
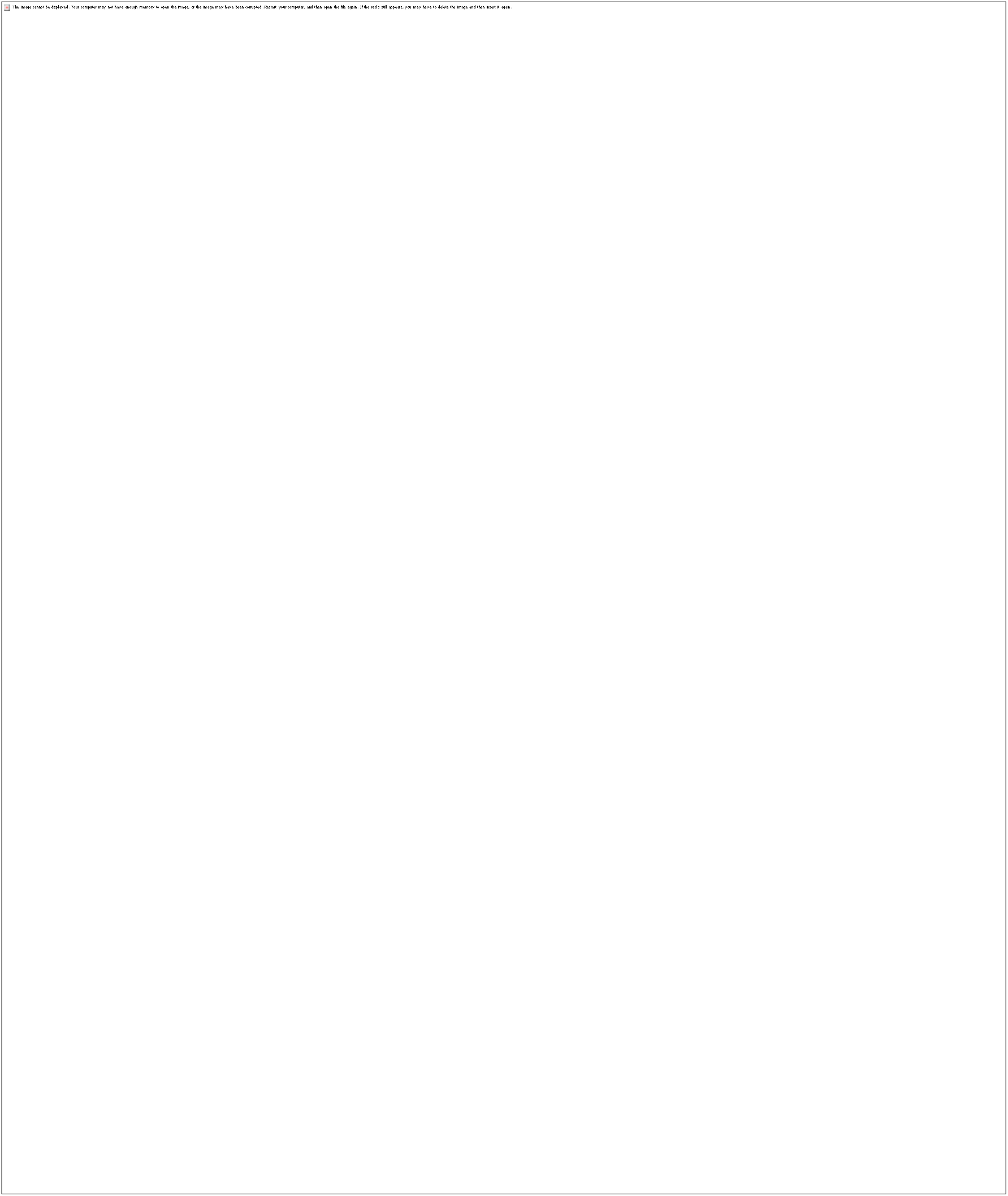
Flowchart for displaying the recruitment process and excluded patients in the study.

## Discussion

nHFO has been shown to be effective and safe when applied to infants with respiratory failure, besides nCPAP or other NIV modes ^19,20^. Our study applied another strategy of using nHFO for infants with respiratory failure who have failed other NIV modes and indicated intubation to prevent complications caused by invasive mechanical ventilation ^11^. We observed the overall success rate within 72 hours was 25 out of 32 cases (78.1%) for rescue indications. This result implies that 78.1% of infants in our study avoided intubation within 72 hours using nHFO, a success that might not have been achieved with previous NIV modes such as nCPAP or NIPPV. The rates of infants who avoided intubation until the completion of nHFO treatment were 23 out of 42 cases (54.8%), applicable to both groups. Invasive ventilation-induced lung injury is a major risk factor for BPD ^5,6^. Infants with BPD are at a higher risk of developing complications, including sepsis, pulmonary arterial hypertension, respiratory failure, and death. Long-term problems include an increased risk of hospital readmissions, respiratory infections, asthma-like symptoms, and an increased risk of poor neurodevelopmental outcomes during infancy and childhood ^4^. In this context, our findings indicate an encouraging advantage of nHFO, given the potential for immediate consequences and the long-term sequelae in premature infants associated with the invasive mechanical ventilation mentioned above.

While all patients were followed to transition to another NIV or intubation, a 72-hour success rate is a more appropriate measure of nHFO efficacy, especially in the rescue group. Respiratory support maintaining infant stability for 72 hours should be considered a successful intervention. Beyond this timeframe, many factors like infection or hemodynamically significant PDA may contribute to therapeutic outcomes. Treatment failure beyond 72 hours should be considered within a broader clinical context, acknowledging these multifactorial contributions rather than solely attributed to nHFO limitations.

Consistent with our findings, prior studies showed a promising ability to reduce intubation in infants with respiratory failure. This effect is attributed to the fact that nHFO has a better CO2 removal efficiency than other NIV modes. A meta-analysis involving 1603 patients was conducted in 2022 to evaluate the efficacy of nHFO ^21^. Results showed that nHFO seems to reduce dramatically the reintubation rates within 72 hours compared to NIPPV. More recently, another meta-analysis including 10 RCTs involving 2031 preterm infants also found that nHFO is an effective intervention for decreasing the intubation or reintubation rate compared with NCPAP ^22^. Previously, a study by Mukerji et al. demonstrated the use of nHFO in a population of 79 preterm infants with NIV failure ^11^. Accordingly, the rate of infants avoiding intubation for the whole and rescue groups was 58% and 55%, respectively. Another study comparing nHFO with NCPAP in 30 preterm infants showed that nHFO could effectively remove carbon dioxide ^23^.

Similarly, a study comparing nHFO with BiPAP in 65 preterm infants also pointed out that nHFO can better reduce CO2 retention and decrease the risk of having apnea ^8^. In a randomized controlled trial comparing nHFO with NCPAP as extubated respiratory support in 206 preterm infants, the nHFO group had significantly lower CO2 levels as well as reintubated rate ^24^. Also, in a meta-analysis that included 1770 patients separated into two groups of nHFO and NIPPV, the CO2 level in the nHFO group was lower than that in the remaining group ^25^.

Our analysis revealed significant changes in pCO_2_ and pH within the 72-hour success group after just one hour of nHFO treatment, which was absent in the 72-hour failure group. We found that nHFO had a good effect not only on improving blood oxygenation but also on improving ventilation and reducing pCO2. These improvements were associated with the success or failure of nHFO. We propose that the absence of early improvement in blood gas parameters following one hour of nHFO initiation may be a prognostic factor for the failure of nHFO. This result raises the question of whether earlier intubation should be performed in this group of children rather than waiting for other clinical signs.

While no significant differences were noted in the birth weight, weight at study entry, and postmenstrual age at study entry between the success and failure groups, the success group of infants appeared to have greater weight and postmenstrual age than the failure group at the time of intervention (995 grams vs. 860 grams, p = 0.056), (29.3 weeks vs. 28.5 weeks, p = 0.123). This finding aligns with the study of Mukerji et al. (865 grams vs. 770 grams, p = 0.284) and (28 weeks vs. 27 weeks, p = 0.252) ^11^. Given the observed trend, we hypothesize that infants with lower weight (≤ 800 grams) and lower postmenstrual age (≤ 27 weeks) may be prognostic factors for nHFO failure. However, these findings require validation in future studies with larger cohorts or via a pooled analysis of available evidence. It is worth mentioning that the rate of surfactant therapy was significantly higher in the failure group compared to the success group, both at 72 hours after nHFO use (100% vs. 73.3%, p<0.001) and upon completion of nHFO use (94.7% vs. 69.6%, p=0.018). This observation aligns with the fact that infants requiring surfactant therapy had poorer lung function, an increased risk of severe bronchopulmonary dysplasia, and a greater need for invasive mechanical ventilation. Additionally, the group of infants who did not require surfactant therapy had a success rate of over 90% in avoiding intubation both at 72 hours and upon completion of nHFO treatment.

Unlike prior investigations comparing nHFO with other NIV modes ^8,10,20^, this study evaluated its efficacy specifically in infants with very severe respiratory failure who had already failed alternative NIV approaches. These infants did not need NIV; rather, they were infants who required intubation if nHFO were unavailable. Thus, our study revealed that the efficacy of nHFO extends beyond a peer-to-peer comparison, indicating its potential superiority over other methods.

The majority of complications related to NIV use were varying degrees of nasal septum injuries. Systemic complications were reported to be rare and accounted for less than 5% of patients ^26^. In fact, abdominal distension was also a common finding in NIV, and it was found to be more severe in nHFO than in other NIV modes due to higher flow and pressure ^26-28^. The feeding intolerance rate in our study was 4.8%, not higher than reported in the literature. In addition, orogastric tube drainage and decreasing flow may help mitigate the problem. Other issues related to safety need to be further examined.

Our study has limitations that need to be declared. First, it is a study that precludes a control group. Therefore, the differences in blood gas parameters after the intervention were not thoroughly evaluated compared to other NIV modes. However, the focus on high-risk infants who had already failed alternative NIV and faced intubation significantly underscores the potential clinical benefit of nHFO in preventing invasive mechanical ventilation in this population. Second, the retrospective design resulted in missing data, limiting a comprehensive assessment of the effect of nHFO on blood gas changes. Finally, inherent variability in clinician judgment to put an infant on nHFO or another respiratory support might lead to an overestimated rating of nHFO efficacy. A prospective study with a larger sample size is needed to evaluate further efficacy, safety, and the setting of appropriate ventilator parameters in this critically ill population.

## Conclusions

Our findings suggest the potential for nHFO as a strategy to reduce invasive mechanical ventilation in high-risk preterm neonates with severe RDS. Future research in this area is warranted, especially on the effects of nHFO on hemodynamics, as well as to optimize ventilator settings and interface selection for optimal safety and efficacy.

## Data Availability

The data presented in this study are available upon reasonable request from the corresponding author. The data contained within this article is not available due to privacy issues.

## Notes

### Competing Interest Statement

The authors have declared no competing interest.

### Funding Statement

The authors received no financial support for the research, authorship, and publication of this article.

### Author Declarations

The study was approved by the Institutional Review Board (IRB) of Children's Hospital 1 (IORG0007285, FWA00009748) on April 7, 2022 (Project No: CS/N1/22/10). A waiver of informed consent was granted by the IRB due to the retrospective nature of the study. Patient data were anonymized and de-identified throughout the analysis.

### Summary of Updates

-Updated the definitions of outcomes more clearly in the manuscript. -Added the statement: The ventilators used in our study do not support synchronization with the patient's breathing during non-invasive ventilation (NIV), in the methods section of the manuscript. -Included a table comparing the blood gas parameters before nHFO use between the 72h success group and 72h failure group (Table 2), and there were no statistically significant differences between these parameters -Added the statement, There were no statistically significant differences in blood gas parameters before the use of nHFO between the 72-hour success group and 72-hour failure group (Table 2), in the Result section. - Included the statement, based on one of the following clinical signs: (1) increased work of breathing such as tachypnea, use of accessory muscles, nasal flaring, grunting, or retractions, (2) hemodynamic instability such as changes in blood pressure and heart rate, or (3) decreased consciousness or lethargy, in the Methods section.

